# A quality framework for perioperative care: rapid review and participatory exercise

**DOI:** 10.1101/2025.08.13.25333583

**Authors:** Kristina Wanyonyi-Kay, Graham Martin, Sarah Ball, Paige Cunnington, Oliver Boney, S. Ramani Moonesinghe, Mary Dixon-Woods, The Perioperative Expert Contributor Group

## Abstract

**Background:** This study addresses the need for a comprehensive, evidence-informed conceptual framework to describe relevant dimensions of quality in perioperative care, moving beyond traditional phase-based models.

**Methods:** We used iterative synthesis of literature as well as consultation and participatory exercises with multidisciplinary professionals and patient representatives, who together formed an Expert Collaborative Group (ECG). The review included primary studies, guidelines, and grey literature identified through searches of PsycINFO, MEDLINE, CINAHL, and institutional websites. The findings were synthesised across studies using an inductive process to build a preliminary framework. Key concepts were identified, mapped, refined, and organised into an initial structure for further development. Fourteen ECG members evaluated a draft version of the framework via the Thiscovery online platform, focusing on relevance and coherence. Their feedback was used to refine the framework, in consultation with clinical experts.

**Results:** Through combination of rapid review of the literature and stakeholder engagement, we were able to develop a framework comprising ten domains that encapsulate structural and process-related features critical to high-quality perioperative care. Feedback from stakeholder exercises was important in improving early versions of the framework, including reordering domains to reflect their centrality and integrating patient and carer perspectives. The resulting perioperative-framework, termed P-Frame, includes domains such as environment and facilities, leadership and governance, organisational culture, shared decision-making, multidisciplinary working, patient optimisation, clinical protocols, post-operative support, staff education, and workforce planning. Importantly, the framework integrates the perspectives of patients and carers, ensuring relevance across clinical, operational, and experiential dimensions of care.

**Conclusions:** P-Frame, based on the available evidence and the perspectives of clinicians and patients, offers a valuable tool for monitoring and improving perioperative care quality as well as supporting research efforts.

**What is already known on this topic:** Perioperative care quality is typically defined within the pre-, intra-, and post-operative phases, restricting the development of broader based quality measurement. This has led to an excess of measures focused on clinical protocols, based on limited evidence and excluding the perspectives of patients and caregivers.

**What this study adds:** This study provides a balanced framework for understanding the domains of quality and safety across all perioperative care settings.

**How this study might affect research, practice, or policy:** It offers a foundation for the development of measures which can be used in audit, research, and targeted improvement initiatives.

## Introduction

While there is no universally agreed definition of quality in health systems, it is generally recognised as a multi-dimensional construct. (1–3) The Institute of Medicine’s (IOM) six key domains of quality – namely safety, effectiveness, patient-centeredness, timeliness, efficiency, and equity - have been of enduring value and influence, (4) but are generic. Understanding what comprises the relevant dimensions of quality and what they look like for specific areas of care is important to quality monitoring, improvement, and research.

In perioperative care, which extends from the initial consideration of surgery through to recovery, a very large number (in excess of 600) of process and structure indicators are published or in use (5, 6) (7) (8) but are typically organised around the temporal phases of care (pre-operative, intra-operative, and post-operative). This phase-based approach risks overlooking important dimensions of quality that cut across timepoints, such as those concerning patient experience and priorities, organisational culture, and care coordination across systems (7). An overarching quality framework is likely to be of benefit to care and to improvement efforts, for example by ensuring clarity about the relevant domains of quality in perioperative care and by supporting balanced and meaningful indicator sets across the domains (8) (9, 10).

To address this need, we aimed to develop a conceptually based framework to identify the quality domains of perioperative care, based on capturing the key concepts set out in the literature in a parsimonious way (4) and on stakeholder engagement and consultation.

## Methods

An overview of the framework development process, which used rapid review of literature and evidence combined with a participatory exercise, is shown in Supplemental Material 1. We sought to ensure our framework would be evidence-informed, contextually relevant, and achievable through a combination of rapid review of the literature and stakeholder engagement. (11, 12) Our approach was guided by Jabareen’s methods for constructing conceptual frameworks (13) and by recommendations for conducting Cochrane Rapid Reviews, including those relating to stakeholder involvement. (14) It involved an iterative process to identify, map, refine, and combine concepts from the literature and then to consult on them.

### Rapid review

The rapid review (PROSPERO registration: CRD42023447192) of existing literature included academic articles from primary research studies, guidelines, standards, and reports and other grey literature sources. Three electronic databases—PsycINFO, MEDLINE, and CINAHL— were searched. Full search strategies are shown in Supplemental Material 2. Articles were screened in two rounds, beginning with titles and abstracts, using an iterative process to refine eligibility criteria and manage the review’s scope. Articles that met the round 2 criteria (Table 1), based on titles and abstracts, were then subject to full-text screening. The grey literature search focused on the websites of key UK-based institutions and agencies. Additional searches were carried out using Google to capture supplementary information. Data were extracted from all included sources using a structured Microsoft Excel-based template, developed specifically for this review. The data extracted included: article details (including article/study type, aims, focus); details on the development and purpose of the domains described; reviewer reflections on study quality and limitations; details on the domains and indicators presented (including domain names and descriptions, sources of domains and how they were conceptualised, and the indicators covered).

**Table 1.** Rapid review eligibility criteria.

|  | **Inclusion criteria** | **Exclusion criteria** |
| --- | --- | --- |
| **Publication type** | Academic and grey literature including:<br>Reports on empirical studies of any design<br>Review articles (including informal reviews)<br>Discussion papers<br>Education pieces/seminars<br>Clinical guidelines and standards<br>Frameworks<br>Grey literature reports<br>Organisational case studies | Commentaries<br>Editorials<br>Perspectives/opinion pieces<br>Letters/correspondence<br>Protocols<br>Clinical case studies |
| **Language** | English | Non-English |
| **Publication date** | 21st July 2013 – 21st July 2023 | Prior to 21st July 2013 |
| **Geographical focus** | Academic literature: Global focus (including low- and middle-income countries). Grey literature: UK focused | Academic literature: None<br>Grey literature: non-UK |
| **Content / topic focus** | **Round 1**<br>Articles that:<br>Relate to the conceptualisation of safe and high-quality perioperative care for patients of any age;<br>Focus on process and structural domains, and any other non-clinical-outcome-based aspects of care;<br>Cover the perspectives of all potential stakeholders (including the views, experiences and satisfaction of service users, carers, and healthcare staff). | **Round 1**<br>Articles that:<br>Relate exclusively to care outside the perioperative period;<br>Focus exclusively on indicators of quality and safety (without broader conceptualisation);<br>Focus exclusively on the clinical outcomes of perioperative care. |
|  | **Round 2** | **Round 2** |
|  | Articles that focus on the topic of conceptualising safe and high-quality care | Articles that describe influences on the safety and/or quality of perioperative care |

### Development of a preliminary conceptual framework

Following data extraction, review findings were synthesised across studies to build a preliminary framework. This inductive process (in line with that proposed by Jabareen) (13) involved the identification and delineation of concepts and constructs identified in the literature, mapping their relationships to one another, combining and dividing them as appropriate, and putting them together into a putative framework for further refinement.

### Participatory exercise

Once we had a preliminary framework, we turned to an online participatory exercise to gather feedback from an Expert Collaborative Group (ECG) consisting of 17 members. This diverse group included perioperative care specialists, anaesthetists, surgeons, nurses, healthcare managers, other healthcare professionals, patient representatives and carers, providing a range of perspectives. Using the Thiscovery online platform, each ECG member reviewed the draft framework (Version 1). They assessed the relevance (i.e. appropriateness in relation to perioperative care quality) of each domain, indicated whether domains should be “kept as is,” “kept but modified,” or “removed,” and provided an overall evaluation of whether the domains collectively constituted an adequate framework for quality in perioperative care. Participants submitted responses via multiple-choice questions with the option to add free-text explanations for their choices.

Ethics approval was given by the Cambridge University Psychology Research Ethics Committee (PRE.2024.045).

### Synthesis and refinement

After collecting feedback, the quantitative and qualitative data from the ECG’s responses were analysed (frequency tables were created to identify the extent of agreement between ECG members regarding whether each domain should be kept as is, modified or removed, and summative content analysis (15) was undertaken to identify common recommendations). The insights gathered were synthesised to inform revisions to the framework, adjusting domains as needed. Where ECG feedback revealed significant discrepancies or disagreements, the research team collaborated with clinical experts to discuss and resolve these differences, refining the framework to ensure its relevance, and coherence.

## Results

Our rapid review and synthesis, combined with the findings of the participatory exercise, enabled the generation of a quality framework which we termed “P-Frame.”

### Rapid review and synthesis

Searches of academic databases returned a total of 3010 academic publications. After removing duplicates, 2403 titles and abstracts of academic papers were screened against inclusion/exclusion criteria. In the first round of title and abstract screening, 1966 records were excluded; subsequently, to manage breadth and volume of evidence, the criteria were revised (Table 1), and titles and abstracts of remaining academic publications were rescreened, excluding an additional 362 records. Full texts of the remaining 75 articles were screened against the revised inclusion/exclusion criteria and a total of 37 academic papers were included in the final review. In addition to academic papers, eight relevant grey literature sources were identified and included in the review. See PRISMA flow diagram (Figure 1) for details.

**Figure 1.**
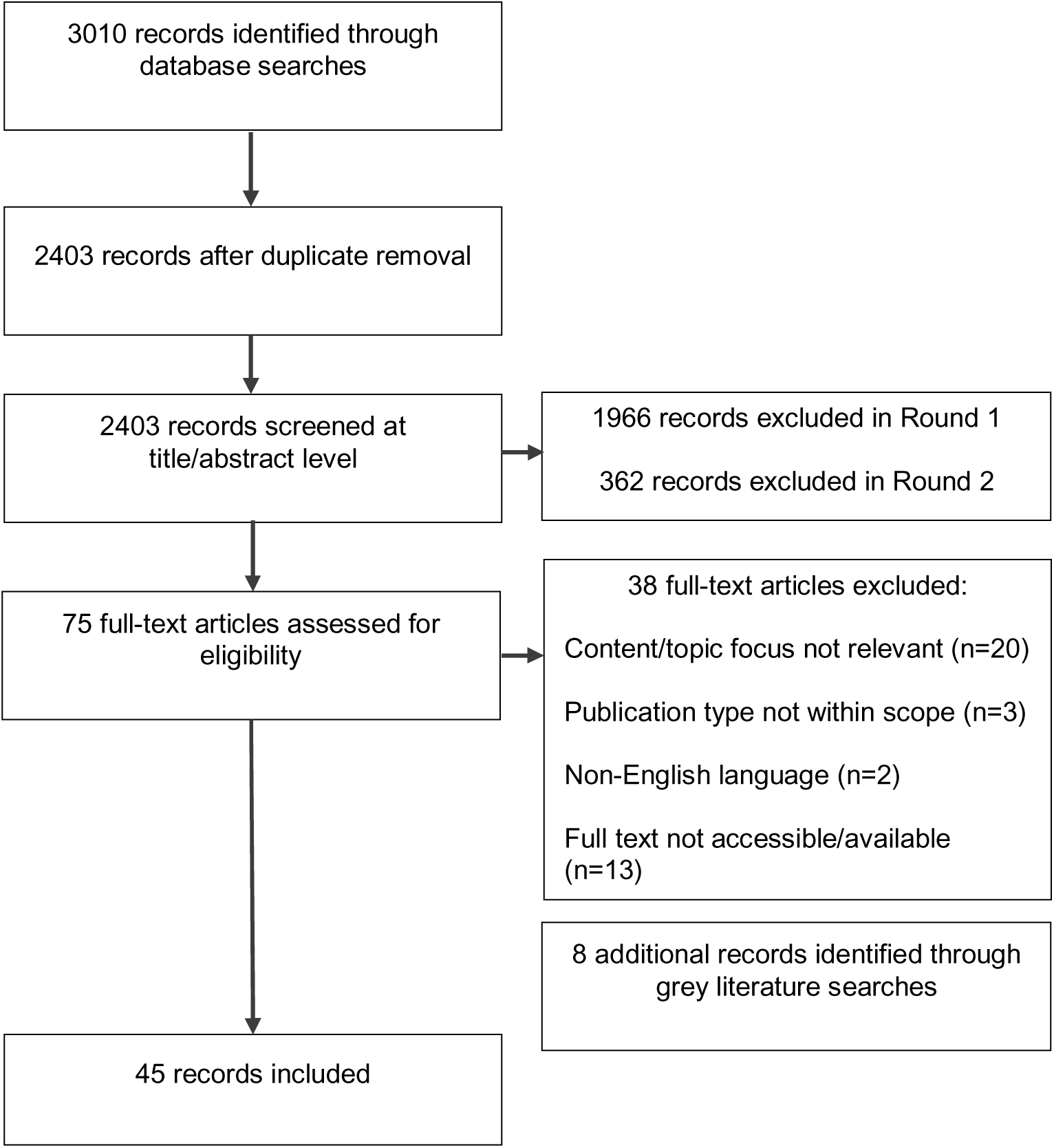
PRISMA Flow Diagram.

### Characteristics of included sources

In total 45 sources were included in the review. Of the 37 academic papers, ten were published between 2013-2015 (16–25), eighteen between 2016 and 2020 (8, 26–42) and the remaining nine after 2021 (43–51). Most academic papers were on the US system (18, 20, 21, 23, 24, 26, 28, 34, 39, 42, 45, 47, 48, 51) or Western Europe (16, 22, 27, 32, 35, 36, 38), including two in the UK (22, 36). Two publications focused on Australia (25, 46), one on Canada (41) and one on Pakistan (33); five had international focus (8, 37, 40, 49, 50) while the remaining six did not explicitly specify geographical focus (17, 19, 29–31, 43, 44).

Fifteen academic publications were reports of empirical studies, including four quantitative (16, 20, 23, 33) and five qualitative studies (22, 35, 36, 38, 47), and six mixed or multimethod studies (18, 24, 32, 37, 40, 46). Eight publications were formal reviews of literature (8, 21, 25, 27, 30, 31, 39, 49), five were discussion or perspective papers (28, 34, 44, 48, 51), six described the development process or characteristics of a framework, indicator set or measurement tool (including a conceptual framework for assessing surgical care quality, patient safety indicators, standards for surgical care of older adults and a measure of post-operative readmission) (17, 19, 26, 29, 43, 50), two reported on quality improvement programmes (42, 45) and one was a consensus statement (41). Six publications focused solely on perioperative care in adult populations (8, 16, 22–24, 49), four considered children (18, 42, 43, 48) and four elderly patients (26, 39, 50, 51). One publication considered both paediatric and adult perioperative care (28); remaining articles did not specify the age-group of interest. Most publications focused on the whole perioperative period (16, 17, 19, 21, 25–30, 32, 39, 40, 43, 44, 49), five considered only postoperative care (22, 34, 36, 37, 46), one the surgery itself (24), and two both surgery and postoperative care (48, 51); remaining studies did not specify their focus within the perioperative care period. Of all papers that explicitly stated the condition or surgery of focus, nine concentrated on general surgery including two focusing on a paediatric population (43, 48), three on cardiac surgeries (24, 30, 51), two on gastrointestinal surgery (16, 19), three on orthopaedic/trauma surgeries (31, 44, 50), and one on plastic surgery (45).

Of the eight grey literature sources, seven were published between 2020 and 2023 (52–58) and one in 2013 (59). All grey literature sources concerned the UK system and consisted of standards (52, 56), guidelines or guidance (53–55, 57, 59) and a report on the Getting it Right First Time (GIRFT) review of Anaesthesia and Perioperative Medicine (58).

### Existing frameworks used in the literature

Within the reviewed literature relating to the conceptualisation of quality in perioperative care, the most frequently referenced model was the IoM model of healthcare quality, (4) which was referred to by nine articles in our sample of the academic literature. (8, 20, 29, 30, 34, 43–45, 48) Another frequently used framework, cited explicitly in four articles (29, 31, 43, 47) and used more widely without explicit reference, was the Donabedian model, with its classic triad of structure, process and outcomes. Neither the IoM nor Donabedian frameworks are specific to the perioperative care context, although they have been used as a means of organising indicators, standards and recommendations relating to the quality of perioperative care – either separately, or in combination with each other (8, 43).

While these were the most commonly cited frameworks, we found that quality indicators, standards and recommendations related to perioperative care were grouped in a variety of other ways within the literature. There was little consistency in the ways in which indicators and standards were subdivided. For example, some were grouped according to broad temporal stages in the perioperative care pathway, along with general aspects or overarching concepts (26, 27, 51). Others were split by treatment topics (such as diagnosis and comorbidity management, discharge planning or pain management) (16, 53) or by clinical versus system-related aspects (25). One article distinguished between different aspects of care, such as between communication with the patient or other providers, patient evaluation, and processes of care (19), while another used groups based on service features such as assurance, reliability and empathy (33). Some articles covered the full range of IoM domains, but others focused on one element of care quality such as patient safety (17, 24, 25, 36, 37) or equity (40, 44), or more narrowly still, on defining quality in surgical palliative care (39) or on the impact of non-technical skills and team working on perioperative care quality (35).

Within the grey literature documents we reviewed, recommendations and quality standards tended to be grouped according to structural categories such as leadership and management (54, 56), workforce, staffing, education, training (54, 55, 57), environment, equipment and facilities (55, 57). The Royal College of Anaesthetists accreditation approach, for example, mapped standards to the quality domains (safe, effective, caring, responsive and well-led) used by the Care Quality Commission (56), the care regulator for England.

### Draft quality framework based on review

Through review of the existing literature described above, we identified a range of *structural* features important to the delivery of high-quality perioperative care including: characteristics of the care environment and facilities; leadership, management and culture; staffing; the systems, pathways and protocols in place; structures to support multidisciplinary working, communication and care co-ordination; specific hospital-level characteristics; and structures to support audit, monitoring and research engagement.

Similarly, a range of *processes* important to perioperative care were identified, including those relating to: patient assessment and documentation; planning and delivery of care; and communication, planning and co-ordination. Further, some aspects of quality might have both structural and process-related features. For example, having a safety protocol in place might be a structural aspect of care, while adherence to the protocol might be process-related. Based on our analysis, we constructed a preliminary draft quality framework (Supplemental Material 3).

### Participatory exercise

Fourteen participants in our ECG, comprising ten healthcare professionals (surgeons, anaesthetists, nurses, healthcare managers and perioperative specialists) and four public contributors, with experience as patients or carers, took part in an online exercise on the online Thiscovery platform to review the draft framework. For each of the domains, respondents indicated: their level of agreement with the statement “This domain encompasses a coherent set of relevant features of quality and safety in perioperative care.” (from 1 – “strongly disagree” to 5 “strongly agree”); and their view on whether the domain should be “kept as is”, “kept but modified” or “removed”. Respondents were also asked “Taken together, do you think these domains make an adequate framework for quality in perioperative care?” indicating “yes”, “no” or “not sure”. Qualitative feedback was provided on the reasons for the responses given.

### Feedback on proposed domains

Overall, a majority of the group agreed that the proposed domains provided a coherent and relevant framework for perioperative care quality. The participants’ level of agreement on whether each domain should be kept, modified or removed and on whether each domain encompasses a coherent set of relevant features are shown in Tables 2 and 3 respectively.

**Table 2.** Level of agreement on whether domains should be kept as is, modified or removed.

| Domain name | Kept as is | Kept but modified | Removed | Not sure | Revised domain name |
| --- | --- | --- | --- | --- | --- |
| Environment & facilities | 8 (57%) | 6 (43%) | 0 (0%) | 0 (0%) | Place, environment, facilities & equipment |
| Leadership & management | 4 (29%) | 10 (71%) | 0 (0%) | 0 (0%) | Leadership, management & governance |
| Organisational culture | 6 (43%) | 6 (43%) | 0 (0%) | 2 (14%) | Organisational culture |
| Staff training & experience | 6 (43%) | 6 (43%) | 2 (14%) | 0 (0%) | Staff education, training & competence |
| Resourcing & staff responsiveness | 7 (50%) | 5 (36%) | 1 (7%) | 1 (7%) | Workforce planning, capacity & support |
| Preoperative care | 4 (29%) | 9 (64%) | 1 (7%) | 0 (0%) | Patient optimisation & preparation: preoperative assessment & care |
| Intraoperative care | 8 (57%) | 6 (43%) | 0 (0%) | 0 (0%) | Clinical care protocols, policies & guidelines |
| Postoperative care | 3 (21%) | 10 (71%) | 0 (0%) | 1 (7%) | Post-operative care, discharge, follow-up & support |
| Continuous monitoring & management | 4 (29%) | 3 (21%) | 2 (14%) | 5 (36%) | N/A |
| Patient communication & shared decision making | 8 (57%) | 5 (36%) | 0 (0%) | 1 (7%) | Shared decision-making & partnership in care |
| Multidisciplinary working & care co-ordination | 7 (50%) | 5 (36%) | 0 (0%) | 2 (14%) | Multidisciplinary working & care co-ordination |

**Table 3.** Level of agreement on relevance of proposed domains.

| Domain name | Strongly agree | Agree | Neither agree nor disagree | Disagree | Strongly disagree | Missing | Revised domain name |
| --- | --- | --- | --- | --- | --- | --- | --- |
| Environment & facilities | 7 (50%) | 7 (50%) | 0 (0%) | 0 (0%) | 0 (0%) | 0 (0%) | Place, environment, facilities & equipment |
| Leadership & management | 5 (36%) | 8 (57%) | 0 (0%) | 0 (0%) | 1 (7%) | 0 (0%) | Leadership, management & governance |
| Organisational culture | 6 (43%) | 4 (29%) | 2 (14%) | 0 (0%) | 1 (7%) | 1 (7%) | Organisational culture |
| Staff training & experience | 4 (29%) | 6 (43%) | 3 (21%) | 1 (7%) | 0 (0%) | 0 (0%) | Staff education, training & competence |
| Resourcing & staff responsiveness | 6 (43%) | 3 (21%) | 2 (14%) | 1 (7%) | 0 (0%) | 2 (14%) | Workforce planning, capacity & support |
| Preoperative care | 4 (29%) | 9 (64%) | 1 (7%) | 0 (0%) | 0 (0%) | 0 (0%) | Patient optimisation & preparation: preoperative assessment & care |
| Intraoperative care | 5 (36%) | 6 (43%) | 0 (0%) | 0 (0%) | 0 (0%) | 3 (21%) | Clinical care protocols, policies & guidelines |
| Postoperative care | 4 (29%) | 3 (21%) | 2 (14%) | 0 (0%) | 1 (7%) | 4 (29%) | Post-operative care, discharge, follow-up & support |
| Continuous monitoring & management | 4 (29%) | 4 (29%) | 1 (7%) | 3 (21%) | 0 (0%) | 2 (14%) | N/A |
| Patient communication & shared decision making | 9 (64%) | 4 (29%) | 0 (0%) | 0 (0%) | 0 (0%) | 1 (7%) | Shared decision-making & partnership in care |
| Multidisciplinary working & care co-ordination | 4 (29%) | 6 (43%) | 0 (0%) | 1 (7%) | 0 (0%) | 3 (21%) | Multidisciplinary working & care co-ordination |

Eight participants considered that the domains collectively formed an adequate framework. All participants agreed on the relevance of four domains. However, free-text feedback raised concerns that the domains originally titled ‘preoperative care,’ ‘intraoperative care,’ and ‘postoperative care’ were overly narrow, lacked balance, and might not resonate with patients. One domain from the initial framework, ‘continuous monitoring and management,’ was deemed redundant, as participants felt that it was insufficiently distinct from the pre-, intra-, and postoperative domains. Consequently, other domains were adapted to incorporate the aspects of quality and safety originally covered by this domain. Additionally, participants recommended reordering the domains to prioritise ‘shared decision-making and partnership in care’ and ‘multidisciplinary working, teamwork, and care coordination’ to better reflect their central role in perioperative care quality.

### Final framework of quality for perioperative care

The feedback from the participatory exercise was used to refine the quality framework including its constituent domains. The final 10-domain framework (“P-Frame”), with definitions and rationale, is shown in Table 4.

**Table 4.** P-Frame: Final quality framework for perioperative care.

| **Domain of perioperative care quality** | **Description and coverage** | **Rationale** |
| --- | --- | --- |
| **Domain 1: Place, environment, facilities and equipment**<br>Appropriate environment, facilities and equipment to support high-quality perioperative care. | This domain focuses on the physical location, environment, facilities and equipment required to enable the provision of high-quality perioperative care. | Initially named 'environment and facilities,' this domain covers the physical setting, equipment, and infrastructure needed for high-quality perioperative care. All participants found it relevant and supported its inclusion in the framework, with 43% suggesting modifications, including renaming it to include "equipment" and "place" and emphasising the need to include in the definition maintenance, patient comfort, and a suitable post-operative home environment.<br>*"I would like to see a reference to [regular] building maintenance included (as well as equipment maintenance). Maintenance is often 'reactive' - emergencies repairs and temporary fixed. I'd prefer to see something much more 'structured' included in this domain"* (Participant 1, patient/carer) |
| **Domain 2: Leadership, management and governance**<br>Appropriate systems of leadership, management and administration to support high-quality perioperative care. | This domain focuses on the leadership, governance, management and administrative structures that need to be in place to support the delivery of high-quality perioperative care and includes a range of predominantly structural indicators. | This domain covers leadership, governance, management, and administrative structures essential for high-quality perioperative care. All participants supported its inclusion in the framework, with 93% agreeing or strongly agreeing that it was relevant. |
|  |  | While the name remained unchanged, 71% recommended modifications, including broadening the focus beyond surgery and anaesthesia, incorporating nursing/therapy leadership, and referencing organisational performance, primary care links, and the triumvirate leadership model.<br>*"I think one important factor is where perioperative care sits as a directorate at trust level - often, anaesthetics and theatres sit together but pre op/ booking and scheduling may sit with the surgical specialties so this really fragments the pathway" (Participant 4, healthcare professional)*<br>*"Where does pre assessment fit? I like to see the triumvirate model; manager, operational lead, clinical lead" (Participant 10, healthcare professional)* |
| **Domain 3: Organisational culture**<br>A culture that promotes a person-centred, improvement-focused, open and inclusive approach to perioperative care. | This domain focuses on the characteristics of organisational culture that support the delivery of high-quality perioperative care. | This domain focuses on a culture that promotes a person-centred, improvement-focused, open and inclusive approach to perioperative care. Most participants (72%) agreed it was relevant, and 86% supported its retention. While the name remained unchanged, suggestions included refining language to include patients and carers and highlighting a learning culture. One participant noted that this domain may be challenging to measure.<br>*"I do agree with this as a domain. But would like to see more mention of carer-centred care in the description. When a* |
|  |  | *carer is involved, it needs to be carer-focused even more than patient-focused. The carers are the ones who have to organise everything and do the work. And they are usually ignored both in terms of pre and post-op planning and in terms of being able to provide feedback. It cannot be a learning culture if you don't learn from carers" (Participant 13, patient/carer)*<br>*"Very wide ranging, hard to measure the overall culture of a hospital/department using simple metrics. There may nominally be structures/ processes for learning, candour, patient-centredness, inclusion, etc - but it's hard to measure how well they translate into the overall culture of an organisation" (Participant 7, healthcare professional)* |
| **Domain 4: Shared decision-making and partnership in care**<br>Shared decision-making and honest, open communication between clinicians and patients/caregivers. | This domain focuses on two-way communication between patients/caregivers and the clinicians providing care, to ensure a common understanding of what perioperative care will involve, and awareness of associated risks and expectations, to enable shared and supported decision-making throughout the perioperative care period. | Originally named 'patient communication and shared decision-making', this domain focuses on two-way communication between patients, caregivers, and clinicians to ensure a common understanding, promote awareness of associated risks and expectations, and support decision-making throughout the perioperative care period. Most participants (93%) agreed on its relevance and the same proportion felt it should be retained in the framework, with 36% suggesting modifications, including reciprocal and tailored communication, patient and carer engagement, and references to advance care planning. |
|  |  | *"This is a very important domain, but need to better capture concept of the patient and family as partners in their care" (Participant 3, patient/carer)*<br>*"I would definitely include the recommendation for [advance] care planning (such as including resuscitation status) in this domain. I would also suggest that we would recommend explaining risk to patients and their families in a meaningful and understandable way for that individual and their family" (Participant 8, healthcare professional)* |
| **Domain 5: Multidisciplinary working and care co-ordination**<br>A multidisciplinary approach to provide joined-up care across all stages of the perioperative pathway, through effective intra- and inter-team communication and care co-ordination. | This domain focuses on multidisciplinary working and care co-ordination, to support the delivery of joined-up care that meets the needs of patients and their caregivers across the whole perioperative care period, from the point of referral and including primary care. | This domain focuses on multidisciplinary working and care co-ordination, to support joined-up care that meets the needs of patients and their caregivers through effective intra- and inter-team communication and care co-ordination. This domain name remained unchanged. Most participants (72%) agreed on its relevance, and 86% supported its retention. While the name remained unchanged, suggestions for changes included broadening the focus beyond clinicians directly involved in perioperative care to include collaboration with various other care providers.<br>*"multi-disciplinary working and care coordination should include effective liaison with Primary Care (e.g. GPs, community nurses etc) and where relevant social care providers"* |
|  |  | (Participant 12, patient/carer) |
| **Domain 6: Patient optimisation and preparation (preoperative assessment and care)**<br>Assessment, optimisation and preparation of the patient during the period between their initial referral for surgery and hospital admission/attendance for their operation. | This domain focuses on the assessment and optimisation of the patient's health and fitness in preparation for surgery - in the time period from their initial referral to their admission to/attendance at hospital for their operation. | Initially named 'preoperative care', this domain focuses on assessing and optimising patients' health before surgery, from referral to hospital admission. Most participants (93%) agreed on its relevance and 93% supported its retention, with 64% suggesting modifications. Feedback emphasised clarifying the preoperative timeline, primary care's role, and incorporating prehabilitation and multidisciplinary team involvement.<br>*"Pre-operative has connotations of admission to induction of anaesthesia, whereas this domain (rightly) is rather wider. Not sure where 'pre-op' is intended to start" (Participant 14, healthcare professional)*<br>*"I do wonder if there is a space in here for something in relation to patient optimisation, the relationship with primary care and overall population health which I feel is becoming a big part of perioperative care. This step would be before preoperative care and could maybe be a title of 'patient preparation, optimisation and coordination'" (Participant 4, healthcare professional)* |
| **Domain 7: Clinical care protocols, policies and guidelines**<br>Availability and use of protocols, policies, guidelines and systems to support clinical care (in the period between the patient's | This domain focuses on the availability and use of protocols, policies, guidelines and systems to support the delivery of high-quality clinical care (in the period between a patient's hospital admission/attendance for their operation and their discharge from the | Initially named 'intraoperative care', this domain covers protocols, guidelines, and systems ensuring high-quality care from hospital admission to postanaesthetic recovery. Most participants (79%) agreed on its relevance and all supported its |
| hospital admission/attendance for their operation – and their discharge from the post-anaesthetic recovery area). | postanaesthetic recovery area). It covers preoperative, intraoperative and immediate-postoperative care provided within this period - including patient assessment, monitoring and documentation, along with the management and delivery of care. | retention, with 43% suggesting modifications. Participants suggested the change to the domain name to differentiate between the phase-specific domains and capture the 'care delivery' component of perioperative care. Furthermore, participants recommended adding references to evidence-based medicine and staffing for safe intraoperative care:<br>*"I think what is intended to be captured is the 'doing stuff' aspect of perioperative care. There isn't a clear distinction for instance about blood management between pre / intra / postop" (Participant 14, healthcare professional)*<br>*"Clarify exactly when intraoperative period starts and finishes" (Participant 3, patient/carer)* |
| **Domain 8: Post-operative care, discharge, follow-up and support**<br>Co-ordinated post-operative care and support from the point at which the patient is discharged from the post-anaesthetic recovery area. | This domain focuses on the activities undertaken after the operation has been completed and the patient has been discharged from the post-anaesthetic recovery area (and the structures in place to support these activities) - to ensure the delivery of high-quality care during this period. It covers care provided in hospital wards, rehabilitation, discharge and post-discharge support. | Initially named 'postoperative care', this domain covers care after surgery, including hospital recovery, rehabilitation, discharge, and post-discharge support. Only 50% of participants agreed on its relevance, but 93% recommended it be retained, with 71% recommending modifications. Suggested modifications included clarifying the postoperative timeframe, considering ward care, and ensuring patient and caregiver communication are foregrounded as part of standard procedures.<br>*"Need to clarify if this is about time in recovery or includes time on the ward. they are different" (Participant 10,* |
|  |  | healthcare professional)<br>*“This domain should include a specific - explicit - reference to SOPs or such like for communication, particularly from the healthcare professional to/with patients, families and carers” (Participant 1, patient/carer)* |
| **Domain 9: Staff education, training, and competence**<br>Education, training and competence of staff across the perioperative pathway, to support the delivery of high-quality, person-centred and pathway-aligned care. | This domain focuses on the education, training and competence that are required by staff in order to deliver high-quality, person-centred and pathway-aligned perioperative care. | Initially named ‘staff training and experience’, this domain focuses on the education and training needed for high-quality, person-centred perioperative care. Most participants (73%) found it relevant, with 86% supporting its retention, half of whom recommended modifications. These included references to multidisciplinary training, inclusive language, and renaming the domain to avoid a potential overlap with workforce planning in the original phrasing.<br>*“Staff need to be given the opportunity and tools to improve and innovate, not just follow protocols and standards. There should be a culture that supports this” (Participant 13, patient/carer)*<br>*“The explanation is too nebulous and does not sufficiently emphasise on the need for patient centred/pathway aligned workforce. Reads very generic and therefore not sure what it adds” (Participant 5, healthcare professional)* |
| **Domain 10: Workforce planning, capacity** | This domain focuses on the staffing required across the perioperative pathway in order to | Initially named ‘resourcing and staff responsiveness’, this domain focuses on |
| **and support**<br>Workforce planning and support to ensure sufficient staffing capacity and responsiveness for the provision of high-quality care. | deliver high-quality perioperative care (taking into consideration the number, availability, and responsiveness of staff). | staffing needs across the perioperative pathway, including staff numbers, availability, and responsiveness. About 64% of participants found it relevant, with 86% supporting its retention, including 36% who recommended modifications. Feedback suggested broadening the scope to include workforce capacity, capability, and planning, and renaming the domain.<br>*“”Workforce” instead of “Staff.”*<br>*Responsiveness sounds like the staff aren’t working properly. Workforce planning is more encompassing”*<br>*(Participant 10, healthcare professional)*<br>*“Very narrow - focussing solely on theatre process, whereas perioperative is wider than that” (Participant 14, healthcare professional)* |

## Discussion

This study expands characterisation of quality in perioperative care beyond the traditional phase-based approach covering preoperative, intraoperative, and postoperative care. Our rapid review confirmed that a conceptual framework specific to perioperative care was lacking, while also indicating that distinctive features of the perioperative pathway limit the usefulness of generic frameworks (7, 8). The structured, collaborative process we used has resulted in a framework (P-Frame) that reflects both evidence from the literature and practical insights from experts and stakeholders in perioperative care, including patients and the public. Its ten domains provide a comprehensive overview of what is important in quality of perioperative care, and will have value in enabling systematic organisation of quality indicators for purposes of monitoring and research, facilitating identification of where indicators may be lacking (such as those relating to patient centredness and equity) (8) and supporting more coherent approaches to assessing and improving care.

The methods we used allowed triangulation of the views of stakeholders with the existing literature, increasing the rigour and face validity of the framework and ensuring consensus on the domains covered. The rapid review was successful in identifying a large number of relevant sources. However, variations in terminology across the literature required interpretive judgment and an iterative synthesis process. For example, where shared decision-making was outlined in the literature, how it was operationalised varied, with some sources constructing it primarily in terms of consent while others took a more encompassing view. These varied views can pose challenges not only at a conceptual level but also in developing reliable quality indicators, which are crucial for meaningful comparisons across settings (60).

The role of stakeholder review of rapid review evidence aligned with Cochrane Rapid Review recommendations, (14) and was important in affirming the relevance of the framework, with unanimous agreement on four domains and minimal disagreement on six others. One domain in the first iteration of the framework—‘continuous monitoring and management’—was dropped following consultation with the ECG, since it was seen as inappropriately isolating aspects of care that were better understood as integral parts of other domains. Some domains benefitted from refined definitions and terminology adjustments. Notably, patient and carer perspectives emphasised the importance of communication and their representation within all domains and subsequent quality measures, helping to address previous concerns about the lack of patient representation in expert panels during indicator development (7, 8). Similarly, stakeholder engagement supported the emergence of culture as a critical domain for perioperative care quality, reflecting Bello et al.’s (61) finding that highlights challenges such as diverse professional groups, dynamic teams, high-stress environments, and conflicting interests. Their work underscores the need for resilient perioperative cultures that can adapt to crises. Similarly, governance was identified as a key pillar of surgical quality (62), alongside the essential role of multidisciplinary collaboration (63).

Several limitations should be considered, however. The rapid review methodology has inherent constraints, including those relating to scope of searches (64) which, in this case, focused on UK-based grey literature. Consequently, some relevant studies might have been excluded, potentially limiting the framework’s applicability across different geographical and health systems. To address this, the participatory exercise engaged a broad, albeit UK-based, expert group to refine and validate the framework, aligning with Cochrane rapid review recommendations (14), which emphasise iterative stakeholder involvement. This approach helped mitigate gaps by integrating diverse expertise including patients and informal carers, ensuring that the framework was both conceptually rigorous and practically applicable.

In conclusion, this study presents a quality framework (P-Frame) informed by stakeholder engagement that is likely to have significant value for perioperative care. Among other things, this framework can inform the review and creation of new indicators, help in ensuring indicator sets are balanced across domains, support clinical audits, drive research initiatives, facilitate the implementation of transferable quality improvement interventions, and ensure that efforts to improve and assure the quality and safety of perioperative care do full justice to the breadth of domains covered by these concepts.

## Statements

### Authors contributions

**KWK:** design, conceptualization, analysis and interpretation of data, writing – first draft, writing – review and editing, critical revision of the manuscript for important intellectual content; **GM**: conceptualization, writing– review and editing, critical revision of the manuscript for important intellectual content; **SB:** data acquisition, analysis and interpretation of data, writing – review and editing, critical revision of the manuscript for important intellectual content; **PC:** writing – review and editing; **OB:** interpretation of data, writing – review and editing; **SRM:** writing – review and editing, critical revision of the manuscript for important intellectual content; **MDW:** funding, conceptualization, writing – first draft, writing – review and editing, critical revision of the manuscript for important intellectual content

## Data Availability

The data are not publicly available as sharing this data could potentially compromise the participant privacy. Data requests can be made to the research team and any such requests will be considered based on data governance.

## Acknowledgements

The authors would like to thank Jo Brand, for supporting with the rapid literature review.

## Contributorship

The Perioperative Expert Contributor Group: Zoe Brummell, Abi Dennington-Price, Jugdeep K Dhesi, Jennifer Dorey, Bob Evans, Michael P W Grocott, Carolyn L Johnston, Emma McCone, Scarlett McNally, Ian Keith Moppett, Zoe Packman, David Selwyn, Ravinder Vohra

## Conflicts of interest

MDW is a founder and non-executive director of THIS Labs, which hosts Thiscovery. She receives no financial benefit from this role. SRM is National Clinical Director for Perioperative and Critical Care at NHS England.

## Funding

This work was funded by the Health Foundation’s grant to the University of Cambridge for The Healthcare Improvement Studies (THIS) Institute (RG88620). THIS Institute is supported by the Health Foundation – an independent charity committed to bringing about better health and health care for people in the UK. The views expressed in this article are those of the authors and not necessarily those of the Health Foundation. The funder played no part in the conduct or writing of this review, or the decision to submit the report for publication.

SRM and OB receive funding from the National Institute for Health and Care Research (NIHR) Central London Patient Safety Research Collaboration; SRM receives funding from the University College London Hospitals NIHR Biomedical Research Centre The views expressed are those of the authors and not necessarily the NIHR or the Department of Health and Social Care.

## References

1. Donabedian A. Evaluating the Quality of Medical Care. The Milbank Memorial Fund Quarterly. 1966;44(3):166–206.

2. Institute of Medicine. Crossing the Quality Chasm: A New Health System for the 21st Century. Washington (DC): Committee on Quality of Health Care in America, National Academies Press (US); 2001.

3. Maxwell RJ. Perspectives In NHS Management: Quality Assessment In Health. British Medical Journal (Clinical Research Edition). 1984;288(6428):1470–2.

4. Institute of Medicine Committee on Quality of Health Care in A. Crossing the Quality Chasm: A New Health System for the 21st Century. Washington (DC): National Academies Press (US). Copyright 2001 by the National Academy of Sciences. All rights reserved.; 2001.

5. CMS. Quality Measures: Centers for Medicare & Medicaid Services; 2018 [Available from: https://www.cms.gov/Medicare/Quality-Initiatives-Patient-Assessment-Instruments/QualityMeasures/index.html.

6. Meyer GS, Nelson EC, Pryor DB, James B, Swensen SJ, Kaplan GS, et al. More quality measures versus measuring what matters: a call for balance and parsimony. BMJ quality & safety. 2012;21(11):964–8.

7. Haller G, Stoelwinder J, Myles PS, McNeil J. Quality and safety indicators in anesthesia: a systematic review. Anesthesiology. 2009;110(5):1158–75.

8. Chazapis M, Gilhooly D, Smith A, Myles P, Haller G, Grocott M, Moonesinghe S. Perioperative structure and process quality and safety indicators: a systematic review. British Journal of Anaesthesia. 2018;120(1):51–66.

9. Perioperative Quality Improvement Programme. PQIP - Perioperative Quality Improvement Programme Report 5 March 2023 to March 2024. 2024.

10. Royal College of Emergency Medicine. RCEM Advisory Statement Regarding the Management of Adults Presenting to the Emergency Department Who May Require an Emergency Laparotomy. 2024.

11. Campbell SM, Roland MO, Buetow SA. Defining quality of care. Social Science & Medicine. 2000;51(11):1611–25.

12. Donabedian A. The quality of care: how can it be assessed? Jama. 1988;260(12):1743–8.

13. Jabareen Y. Building a conceptual framework: philosophy, definitions, and procedure. International journal of qualitative methods. 2009;8(4):49–62.

14. Garritty C, Gartlehner G, Nussbaumer-Streit B, King VJ, Hamel C, Kamel C, et al. Cochrane Rapid Reviews Methods Group offers evidence-informed guidance to conduct rapid reviews. Journal of Clinical Epidemiology. 2021;130:13–22.

15. Hsieh H-F, Shannon SE. Three approaches to qualitative content analysis. Qualitative Health Research. 2005;15(9):1277–88.

16. De Boer M, Ramrattan MA, Boeker EB, Kuks PFM, Boermeester MA, Lie AHL. Quality of pharmaceutical care in surgical patients. PLoS ONE. 2014;9(7):e101573.

17. Emond YE, Stienen JJ, Wollersheim HC, Bloo GJ, Damen J, Westert GP, et al. Development and measurement of perioperative patient safety indicators. British journal of anaesthesia. 2015;114(6):963–72.

18. Espinel AG, Shah RK, Beach MC, Boss EF. What parents say about their child’s surgeon: parent-reported experiences with pediatric surgical physicians. JAMA Otolaryngology-Head & Neck Surgery. 2014;140(5):397–402.

19. Halverson AL, Sellers MM, Bilimoria KY, Hawn MT, Williams MV, McLeod RS, Ko CY. Identification of process measures to reduce postoperative readmission. Journal of gastrointestinal surgery : official journal of the Society for Surgery of the Alimentary Tract. 2014;18(8):1407–15.

20. Kalish BT, Vollmer CM, Kent TS, Nealon WH, Tseng JF, Callery MP. Quality assessment in pancreatic surgery: what might tomorrow require? Journal of gastrointestinal surgery : official journal of the Society for Surgery of the Alimentary Tract. 2013;17(1):86-p.93.

21. Kim FJ, da Silva RD, Gustafson D, Nogueira L, Harlin T, Paul DL. Current issues in patient safety in surgery: A review. Patient Safety in Surgery. 2015;9(1):26.

22. Pucher PH, Aggarwal R, Singh P, Tahir M, Darzi A. Identifying quality markers and improvement measures for ward-based surgical care: a semistructured interview study. American journal of surgery. 2015;210(2):211–8.

23. Sacks GD, Lawson EH, Dawes AJ, Russell MM, Maggard-Gibbons M, Zingmond DS, Ko CY. Relationship Between Hospital Performance on a Patient Satisfaction Survey and Surgical Quality. JAMA surgery. 2015;150(9):858–64.

24. Thompson DA, Marsteller JA, Pronovost PJ, Gurses A, Lubomski LH, Goeschel CA, et al. Locating Errors Through Networked Surveillance: A Multimethod Approach to Peer Assessment, Hazard Identification, and Prioritization of Patient Safety Efforts in Cardiac Surgery. Journal of patient safety. 2015;11(3):143–51.

25. Wacker J, Staender S. The role of the anesthesiologist in perioperative patient safety. Current Opinion in Anaesthesiology. 2014;27(6):649–56.

26. Berian JR, Rosenthal RA, Baker TL, Coleman J, Finlayson E, Katlic MR, et al. Hospital Standards to Promote Optimal Surgical Care of the Older Adult: A Report From the Coalition for Quality in Geriatric Surgery. Annals of surgery. 2018;267(2):280–90.

27. Bresadola V, Biddau C, Puggioni A, Tel A, Robiony M, Hodgkinson J, Leo CA. General surgery and COVID-19: review of practical recommendations in the first pandemic phase. Surgery today. 2020;50(10):1159–67.

28. Calabro KA, Raval MV, Rothstein DH. Importance of patient and family satisfaction in perioperative care. Seminars in pediatric surgery. 2018;27(2):114–20.

29. Citron I, Saluja S, Amundson J, Ferreira RV, Ljungman D, Alonso N, et al. Surgical quality indicators in low-resource settings: A new evidence-based tool. Surgery (United States). 2018;164(5):946–52.

30. Coulson TG, Mullany DV, Reid CM, Bailey M, Pilcher D. Measuring the quality of perioperative care in cardiac surgery. European Heart Journal - Quality of Care and Clinical Outcomes. 2017;3(1):11–9.

31. DeBaun MRMD, Chen MJMD, Bishop JAMD, Gardner MJMD, Kamal RNMD, DeBaun MR, et al. Orthopaedic Trauma Quality Measures for Value-Based Health Care Delivery: A Systematic Review. Journal of Orthopaedic Trauma. 2019;33(2):104–10.

32. Egbert N, Babitsch B, Hübner U. Learning from CIRS to optimise patient safety in handovers. GMS Medizinische Informatik, Biometrie und Epidemiologie. 2020;16(3):1–15.

33. Fatima I, Humayun A, Anwar MI, Iftikhar A, Aslam M, Shafiq M. How Do Patients Perceive and Expect Quality of Surgery, Diagnostics, and Emergency Services in Tertiary Care Hospitals? An Evidence of Gap Analysis From Pakistan. Oman Medical Journal. 2017;32(4):297–305.

34. Fleisher LA. Quality Anesthesia: Medicine Measures, Patients Decide. Anesthesiology. 2018;129(6):1063–9.

35. Hanssen I, Smith Jacobsen IL, Skramm SH. Non-technical skills in operating room nursing: Ethical aspects. Nursing ethics. 2020;27(5):1364–72.

36. Hassen Y, Singh P, Pucher PH, Johnston MJ, Darzi A. Identifying quality markers of a safe surgical ward: An interview study of patients, clinical staff, and administrators. Surgery. 2018;163(6):1226–33.

37. Hassen YAM, Johnston MJ, Singh P, Pucher PH, Darzi A. Key Components of the Safe Surgical Ward: International Delphi Consensus Study to Identify Factors for Quality Assessment and Service Improvement. Annals of surgery. 2019;269(6):1064–72.

38. Jangland E, Nyberg B, Yngman-Uhlin P. ’It’s a matter of patient safety’: understanding challenges in everyday clinical practice for achieving good care on the surgical ward - a qualitative study. Scandinavian Journal of Caring Sciences. 2017;31(2):323–31.

39. Lee KC, Senglaub SS, Walling AM, Mosenthal AC, Cooper Z. Quality Measures in Surgical Palliative Care: Adapting Existing Palliative Care Measures to Improve Care for Seriously Ill Surgical Patients. Annals of surgery. 2019;269(4):607–9.

40. Santhirapala V, Peden CJ, Meara JG, Biccard BM, Gelb AW, Johnson WD, et al. Towards high-quality peri-operative care: a global perspective. Anaesthesia. 2020;75:e18–e27.

41. Urbach DR, Karimuddin AA, Wei A, Zabolotny BP, Lefebvre G, Walsh M, et al. A Canadian strategy for surgical quality improvement. Canadian Journal of Surgery. 2019;62(6):E16–E8.

42. Wang KS, Cummings J, Stark A, Houck C, Oldham K, Grant C. Optimizing Resources in Children’s Surgical Care: An Update on the American College of Surgeons’ Verification Program. Pediatrics. 2020;145(5):1–7.

43. He K, Cramm SL, Rangel SJ. Defining high-quality care in pediatric surgery: Implications for performance measurement and prioritization of quality and process improvement efforts. Seminars in pediatric surgery. 2023;32(2):151274.

44. Knowlton LM, Zakrison T, Kao LS, McCrum ML, Agarwal S, Bruns B, et al. Quality care is equitable care: a call to action to link quality to achieving health equity within acute care surgery. Trauma Surgery and Acute Care Open. 2023;8(1):e001098.

45. Manahan MA, Aston JW, Bello RJ, Siotos C, Demski R, Cooney CM, et al. Establishing a Culture of Patient Safety, Quality, and Service in Plastic Surgery: Integrating the Fractal Model. Journal of patient safety. 2021;17(8):e1553–e8.

46. McGuire L, Schultz TJ, Kelly J. Developing a Model of Care for a 4-to 6-Bedded Postanesthetic Recovery Unit: A Delphi Study. Journal of perianesthesia nursing : official journal of the American Society of PeriAnesthesia Nurses. 2021;36(4):398–405.

47. Mull HJ, Rosen AK, Charns MP, Itani KMF, Rivard PE. Identifying Risks and Opportunities in Outpatient Surgical Patient Safety: A Qualitative Analysis of Veterans Health Administration Staff Perceptions. Journal of patient safety. 2021;17(3):e177–e85.

48. Olbrecht VA, Uffman JC, Morse RB, Engelhardt T, Tobias JD. Setting a universal standard: Should we benchmark quality outcomes for pediatric anesthesia care? Paediatric anaesthesia. 2022;32(8):892–8.

49. Pinto J, Matias AC, Sá L, Amaral A. Quality Indicators in Ambulatory Care Surgery: A Scoping Review. Nursing Economics. 2022;40(5):215–29.

50. Sermon A, Slock C, Coeckelberghs E, Seys D, Panella M, Bruyneel L, et al. Quality indicators in the treatment of geriatric hip fractures: literature review and expert consensus. Archives of osteoporosis. 2021;16(1):152.

51. Zhongmin L, Shirakawa N, Chen A, Fu-Hai J, Hong L. Methodology in coronary artery bypass surgery quality assessment. Journal of Geriatric Cardiology. 2022;19(11):867–75.

52. Centre for Perioperative Care. National Safety Standards for Invasive Procedures 2 (NatSSIPs): short version. 2023.

53. National Institute for Health and Care Excellence. Perioperative care in adults. 2020.

54. Royal College of Anaesthetists. Chapter 1: Guidelines for the Provision of Anaesthesia Services: The Good Department 2023 2023 [Available from: https://www.rcoa.ac.uk/gpas/chapter-1.

55. Royal College of Anaesthetists. Chapter 2: Guidelines for the Provision of Anaesthesia Services for the Perioperative Care of Elective and Urgent Care Patients 2023 2023 [Available from: https://www.rcoa.ac.uk/gpas/chapter-2.

56. Royal College of Anaesthetists Anaesthesia CSA. Accreditation standards 2023. 2023.

57. Royal College of Nursing. Day Surgery for Children and Young People. 2020.

58. Dr Chris Snowden, Dr Mike Swart. Anaesthesia and Perioperative Medicine GIRFT Programme National Speciality Report. 2021.

59. Royal College of Surgeons. Good Surgical Practice. 2013.

60. Bottle A, Kristensen PK. Variation in quality of care between hospitals: how to identify learning opportunities. BMJ Quality & Safety. 2024;33(7):413.

61. Bello C, Filipovic MG, Andereggen L, Heidegger T, Urman RD, Luedi MM. Building a well-balanced culture in the perioperative setting. Best Practice & Research Clinical Anaesthesiology. 2022;36(2):247–56.

62. Royal College of Surgeons of England. Clinical Governance of the Surgical Care Team 2025 [Available from: https://www.rcseng.ac.uk/standards-and-research/standards-and-guidance/service-standards/surgical-care-team-guidance/clinical-governance/.

63. Taberna M, Gil Moncayo F, Jané-Salas E, Antonio M, Arribas L, Vilajosana E, et al. The Multidisciplinary Team (MDT) Approach and Quality of Care. Front Oncol. 2020;10:85.

64. King VJ, Stevens A, Nussbaumer-Streit B, Kamel C, Garritty C. Paper 2: Performing rapid reviews. Systematic Reviews. 2022;11(1):151.

