## Supplemental material for "A quality framework for perioperative care: rapid review and participatory exercise"

**Supplemental material 1**. Flow diagram of the framework development process

**1. Rapid literature review**

Perioperative care stakeholders (N=17) invited to take part in an online participatory exercise to review the draft conceptual framework

Each domain in the framework assessed for relevance in relation to perioperative care quality

Overall framework evaluated to determine whether the domains collectively constitute an adequate framework of quality in perioperative care

Feedback from stakeholders analysed using qualitative analysis (summative content analysis) and quantitative analysis (based on frequency tables) and synthesised to inform revisions to the framework

PsycINFO, MEDLINE and CINAHL searched to identify relevant academic articles and grey literature

Identified articles screened against eligibility criteria. Information about constituent elements of high-quality perioperative care extracted from included articles using a purpose-designed structured Excel template

Key concepts and constructs of high-quality perioperative care synthesised, mapped, refined and combined into putative frameworks across multiple stages

Preliminary conceptual framework of high-quality perioperative care developed

**3. Stakeholder review of draft conceptual framework**

**Final quality framework (P-Frame) for perioperative care**

**2. Development of draft conceptual framework**

N=14 perioperative care stakeholders took part in the online participatory exercise

**Supplemental material 2**. Search strategies for the rapid literature review

| Search string | Database or further sources | Results | Date |
| --- | --- | --- | --- |
| "quality of health care"/ or process assessment, health care/ or quality assurance, health care/ or quality improvement/ or risk adjustment/ or "standard of care"/ OR patient harm/ or patient safety/ or safety management/ AND   perioperative care/ or intraoperative care/ or postoperative care/ or preoperative care/ OR perioperative period/ or intraoperative period/ or postoperative period/ or preoperative period/ OR (pre-op* or preop* or pre-op* or postop* or post-op* or postsurg* or post-surg* or presurg* or pre-surg* or intraop* or intra-op* or anaesth* or anesth* or preanaesth* or pre-anaesth* or postanaesth* or post-anaesth* or preanesth* or pre-anesth* or postanesth* or post-anesth* or surg* or operat*) adj2 (care or period or system* or process* or safety or complication* or emergenc* or adverse or recover* or mortality or morbidity or pain or experienc*)).ti. or ((periop* or peri-op* or preop* or pre-op* or postop* or post-op* or postsurg* or post-surg* or presurg* or pre-surg* or intraop* or intra-op* or anaesth* or anesth* or preanaesth* or pre-anaesth* or postanaesth* or post-anaesth* or preanesth* or pre-anesth* or postanesth* or post-anesth* or surg* or operat*) adj2 (care or period or system* or process* or safety or complication* or emergenc* or adverse or recover* or mortality or morbidity or pain or experienc*)).ab. AND  ((quality or safe* or process* or structure*) adj2 (domain* or measur* or assess* or criter* or standard* or concept* or idea* or feature* or component* or element* or constitu* or dimension* or barrier* or facilitat*)).ti. or ((quality or safe* or process* or structure*) adj2 (domain* or measur* or assess* or criter* or standard* or concept* or idea* or feature* or component* or element* or constitu* or dimension* or barrier* or facilitat*)).ab. | Medline | 768 | 2023-07-21 |
| health care quality/ OR patient safety/ or patient harm/ or patient risk/ AND  perioperative care/ or perioperative period/ or peroperative care/ or postanesthesia care/ or postoperative care/ OR ((periop* or peri-op* or preop* or pre-op* or postop* or post-op* or postsurg* or post-surg* or presurg* or pre-surg* or intraop* or intra-op* or anaesth* or anesth* or preanaesth* or pre-anaesth* or postanaesth* or post-anaesth* or preanesth* or pre-anesth* or postanesth* or post-anesth* or surg* or operat*) adj2 (care or period or system* or process* or safety or complication* or emergenc* or adverse or recover* or mortality or morbidity or pain or experienc*)).ti. or ((periop* or peri-op* or preop* or pre-op* or postop* or post-op* or postsurg* or post-surg* or presurg* or pre-surg* or intraop* or intra-op* or anaesth* or anesth* or preanaesth* or pre-anaesth* or postanaesth* or post-anaesth* or preanesth* or pre-anesth* or postanesth* or post-anesth* or surg* or operat*) adj2 (care or period or system* or process* or safety or complication* or emergenc* or adverse or recover* or mortality or morbidity or pain or experienc*)).ab. AND  ((quality or safe* or process* or structure*) adj2 (domain* or measur* or assess* or criter* or standard* or concept* or idea* or feature* or component* or element* or constitu* or dimension* or barrier* or facilitat*)).ti. or ((quality or safe* or process* or structure*) adj2 (domain* or measur* or assess* or criter* or standard* or concept* or idea* or feature* or component* or element* or constitu* or dimension* or barrier* or facilitat*)).ab. | Embase | 1703 | 2023-07-21 |
| (MM "Accountability") OR (MM "Quality of Health Care") OR (MM "Quality Improvement") OR (MM "Quality Assessment") OR (MM "Quality Assurance") OR (MM "Quality Management, Organizational") OR (MM "Patient Safety") OR (MM "Adverse Health Care Event") OR (MM "Risk Management") OR (MM "Incident Reports") AND  (MH "Perioperative Care+") OR (MH "Intraoperative Care+") OR (MH "Postoperative Care+") OR (MH "Preoperative Care+") OR (MM "Preoperative Period") OR (MM "Intraoperative Period") OR (MM "Surgery, Operative") OR TI ( ((periop* or peri-op* or preop* or pre-op* or postop* or post-op* or postsurg* or post-surg* or presurg* or pre-surg* or intraop* or intra-op* or anaesth* or anesth* or preanaesth* or pre-anaesth* or postanaesth* or post-anaesth* or preanesth* or pre-anesth* or postanesth* or post-anesth* or surg* or operat*) N2 (care or period or system* or process* or safety or complication* or emergenc* or adverse or recover* or mortality or morbidity or pain or experienc*)) ) OR AB ( ((periop* or peri-op* or preop* or pre-op* or postop* or post-op* or postsurg* or post-surg* or presurg* or pre-surg* or intraop* or intra-op* or anaesth* or anesth* or preanaesth* or pre-anaesth* or postanaesth* or post-anaesth* or preanesth* or pre-anesth* or postanesth* or post-anesth* or surg* or operat*) N2 (care or period or system* or process* or safety or complication* or emergenc* or adverse or recover* or mortality or morbidity or pain or experienc*)) ) AND  TI ( ((quality or safe* or process* or structure*) N2 (domain* or measur* or assess* or criter* or standard* or concept* or idea* or feature* or component* or element* or constitu* or dimension* or barrier* or facilitat*)) ) OR AB ( ((quality or safe* or process* or structure*) N2 (domain* or measur* or assess* or criter* or standard* or concept* or idea* or feature* or component* or element* or constitu* or dimension* or barrier* or facilitat*)) ) | CINAHL | 539 | 2023-07-21 |

**Supplemental material 3**. Preliminary draft conceptual framework

| **Domain of perioperative care quality** | **Description and coverage** |
| --- | --- |
| **1. Environment and facilities**  Appropriate and accessible environment and facilities for perioperative care provision. | This domain focuses on the environment and facilities required to enable the provision of safe and high-quality perioperative care and includes predominantly structural indicators.  Structural indicators within this domain could include those relating to:   - **the availability and capacity of appropriate facilities and care environments for perioperative care** (in line with relevant guidelines and differentiated according to patient needs). - **the availability of appropriate and well-maintained equipment** (and maintenance schedules/protocols to support this). - **access to and safe/appropriate storage of medicines and medical supplies.** - **the layout, physical characteristics, maintenance and cleanliness of care environments.** - **the aesthetics and atmosphere of these environments.** - **geographical location and travel infrastructure** (and whether this supports manageable and timely travel for patient and staff access).   Process indicators could include those relating to:   - **adherence to equipment maintenance protocols** (and accurate documentation of this). |
| **2. Leadership and management**  Leadership, governance, management and administration to support quality perioperative care. | This domain focuses on the leadership, governance, management and administrative structures that need to be in place to support the delivery of safe and high-quality perioperative care and includes a range of structural indicators.  Structural indicators within this domain could include those relating to:   - **planning and organisational strategy** (such as, whether there is a live/annually reviewed operational plan). - **the existence of leadership/governance structures** with clearly defined roles and responsibilities. - **appropriate clinical leadership across emergency and elective surgery and anaesthesia** (to ensure best practice is followed, foster a learning culture and champion change). - **management, administration and co-ordination of services** (such as, whether there is collaboration between clinical, managerial and administrative services to facilitate an efficient patient journey and the availability of appropriate administrative and secretarial support). |
| **3. Organisational culture**  High quality care culture (open, learning, patient-centred and inclusive culture). | This domain focuses on the characteristics of organisational culture that support the delivery of safe and high-quality perioperative care and includes a range of predominantly structural indicators.  Structural indicators within this domain could include those relating to:   - **learning culture** (such as, whether there is a focus on, and structures and processes are in place to support, quality and safety improvement - by learning from adverse events, near misses and feedback). - **candour/openness** (such as, whether staff have the opportunity to speak up without fear of blame). - **patient-centred culture** (such as whether patient experience and feedback are used to drive improvement in the quality and safety of perioperative care, and staff have the capacity, capability, willingness and support to deliver patient-centred care). *[Note: indicators could include those based on direct patient experience]* - **culture of inclusion** (such as, whether equality, diversity and inclusion are considered in relation to recruitment, training, and workforce characteristics as well as service provision, and reflected in local policies on provision of care tailored to individual patient needs and/or protected characteristics). *[Note: indicators could include those based on direct patient experience in relation to equity]*   Process indicators within this domain could include those relating to:   - **adherence to policies and protocols to ensure the provision of timely and appropriate perioperative care to all patients**, regardless of their personal characteristics and circumstances (such as, whether there are equal levels of adherence to standardized protocols between patients with different characteristics and whether steps are taken to ensure the needs of patients with specific risks or requirements are considered and addressed at all stages of perioperative care delivery). |
| **4. Staff skills and experiences**  Staff professional skills, training and experience. | This domain focuses on the professional skills, training and experience that are required by staff in order to deliver safe and high-quality perioperative care and includes a range of structural indicators.  Structural indicators within this domain could include those relating to:   - **appropriate education, training and experience** (such as, whether policies are in place on requirements for education and training, whether staff at all levels have appropriate training and experience in line with national standards, and the status of the hospital in relation to staff professional development, training and experience - e.g. sufficiently high surgical case volume, accreditation status and having teaching hospital status). - **professional competence** (such as, whether staff are aware of best practice protocols, technical and non-technical skills to ensure the delivery of high-quality and safe perioperative care, staff maintaining competence in all areas of their practice). |
| **5. Resourcing and staff responsiveness**  Staffing (resourcing, availability and responsiveness). | This domain focuses on the staffing required in order to deliver safe and high-quality perioperative care (taking into consideration the number, availability and responsiveness of staff) and includes a range of predominantly structural indicators.  Structural indicators within this domain could include those relating to:   - **appropriate resourcing and skill mix** (such as, whether there is an appropriate number of skilled staff/clinicians to support the provision of safe, effective, efficient and timely care, including specialist teams as required, with capacity to support emergency cases and complex patients, and whether appropriate levels of supervision are in place). - **staff availability, flexibility and responsiveness** (such as whether the required staff are available when needed, there is sufficient flexibility in staffing to enable to provision of timely care, resource allocation/rota management processes are in place that are compliant with national guidelines, policies for 24h cover of emergency surgeries and prioritising emergency cases based on clinical urgency and staff awareness of emergency call systems and escalation processes). - **support to ensure availability/responsiveness of staff** (such as, whether there are facilities for rest and refreshment for staff on- call, opportunities to facilitate staff development and retention, support for the health and wellbeing of staff members, and workload monitoring in order to plan future capacity).   Process indicators within this domain could include those relating to:   - **adherence to protocols and guidelines to ensure that appropriately trained and experienced staff are present - for operative procedures, in transfer and in recovery.** |
| **6. Preoperative care**  Preoperative assessment, monitoring, documentation and care delivery. | This domain focuses on the activities undertaken within the preoperative period (and structures in place to support these activities) to ensure the delivery of safe and high-quality care. It covers preoperative patient assessment, monitoring and documentation, along with the management and delivery of safe and high-quality care. It includes a variety of both structural and process indicators.  Structural indicators within this domain could include those relating to:   - **the protocols, policies and systems in place to support preoperative patient assessments, investigations, monitoring and recording-keeping** (such as, whether there are protocols in place for preoperative patient assessment, accurate documentation, and prospective risk analysis) and preoperative care management and delivery (such as, whether there are policies on the use of preoperative prophylactic therapies/ preparation, or management strategies to avoid delays or cancellations of surgeries).   Process indicators within this domain could include those relating to:   - **adherence to protocols, policies and guidelines for preoperative patient assessment, risk analysis, monitoring, review and documentation, and for preoperative care management and delivery.** - **use of available preoperative checklists/tools intended to support adherence to guidelines or measures to avoid errors.** An example would be WHO surgery checklist. |
| **7. Intraoperative care**  Intraoperative assessment, monitoring, documentation and care-delivery. | This domain focuses on the activities undertaken within the intraoperative period (and structures in place to support these activities) to ensure the delivery of safe and high-quality care. It covers intraoperative patient assessment, monitoring and documentation, along with the management and delivery of safe and high-quality care in the intraoperative period. It includes a variety of both structural and process indicators.  Structural indicators within this domain could include those relating to:   - **the protocols, policies and systems in place to support intraoperative patient assessments, investigations, monitoring and recording-keeping; and intraoperative care management and delivery** (such as, whether there are protocols for dealing with complications and providing emergency care, or guidelines on medication, anaesthetic care or surgical procedures and blood management)   Process indicators within this domain could include those relating to:   - **adherence to protocols, policies and guidelines for intraoperative patient assessment, monitoring, review and documentation and for intraoperative care management and delivery** - **intraoperative use of available checklists/tools intended to support adherence to guidelines or measures to avoid errors** (such as, surgical safety checklists, procedure specific checklists, measures to ensure proper positioning on the operating table/correct site for surgery). |
| **8. Postoperative care**  Postoperative assessment, monitoring, documentation, care delivery, discharge and follow-up. | This domain focuses on the activities undertaken within the postoperative period (and structures in place to support these activities) to ensure the delivery of safe and high-quality care. It covers postoperative patient assessment, monitoring and documentation, along with the management and delivery of safe and high-quality care in the postoperative period. It includes a variety of both structural and process indicators.  Structural indicators within this domain could include those relating to:   - **the protocols, policies and systems in place to support postoperative patient assessments, investigations, monitoring and recording-keeping and to guide postoperative care management and delivery** (such as, whether there are protocols and policies for postoperative management of pain, nausea and vomiting, optimal discharge policies and protocols for medication management). - **the availability of timely surgical follow-up** (such as, follow-up available within 30 days following hospital discharge).   Process indicators within this domain could include those relating to:   - **adherence to protocols, policies and guidelines, to ensure that the postoperative care provided is safe and of high quality** and that it is provided in a timely and efficient manner. - **use of available checklists/tools intended to support adherence to guidelines** (such as, a checklist or protocol for direct transfer of care from procedure room to intensive care) - **administrative measures** (such as rates of errors/adverse events/delays, length of stay in hospital, readmission rates, costs of care, complications, or readmissions). |
| **9. Continuous perioperative monitoring and management**  Continuous monitoring, blood and medication management and maintenance of vital signs. | This domain focuses on the continuous patient monitoring and management activities undertaken across the whole perioperative period (and structures in place to support these activities) to ensure the delivery of safe and high-quality care. It covers continuous monitoring and documentation across all phases of care, blood and medication management and maintenance of vital signs. It includes a variety of both structural and process indicators.  Structural indicators within this domain could include those relating to:   - **the protocols, policies and systems in place to support continuous monitoring of patients and management of their condition** (such as, whether there are agreed protocols and policies for continuous assessment, and for medication and blood management – including standard protocols for use of anticoagulants or antibiotics, guidelines on intravenous fluid therapy and enhanced recovery protocols).   Process indicators within this domain could include those relating to:   - **adherence to protocols, policies and guidelines, to ensure that patients are monitored and have their condition managed appropriately throughout the perioperative period.** |
| **10. Patient communication and shared decision making**  Shared decision making and communication with patients and caregivers. | This domain focuses on communication with patients and caregivers to ensure that they understand what their perioperative care will involve, and are aware of associated risks and expectations, to enable their involvement in shared decision making throughout the perioperative care period. It includes a variety of both structural and process indicators.  Structural indicators within this domain could include those relating to:   - **the structures and processes in place to ensure the availability of advice, guidance and support for patients and caregivers** (such as, to support the provision of patient information in accessible formats, access to specialist advice that is tailored to individual needs, preoperative discussions between clinicians and patients/caregivers to ensure a shared understanding about care). - **the policies, guidance and systems in place to ensure respect for the wishes of patients and caregivers** (such as, the existence of a trust/board resuscitation policy with specific reference to the perioperative period, guidance on implementing advanced care plans in the perioperative period.)   Process indicators within this domain could include those relating to:   - **provision of timely, sufficient, expert and appropriately tailored information and support to patients and caregivers throughout the perioperative period** (such as, whether information is provided on *when* procedures will be performed and *what* the intended treatment will involve, whether *postoperative/discharge instructions* are provided, whether communication is consultant-led, and clear and compassionate, whether support is provided for families and caregivers in the hospital environment and prior to discharge, including education on their role in pain management). - **provision of support for and evidence of patient involvement in shared decision making** (such as, whether steps are taken to ensure patients and their family/caregivers are engaged in shared decision-making – including in relation to discharge planning, whether patients themselves report feeling involved in decision making in relation to their care). *[Note: indicators could include those based on patient reported outcome measures such as patient activation/ability to self-manage condition]* - **respect for the wishes of patients and caregivers** (such as, whether end of life discussions are undertaken where appropriate, informed consent is taken for anaesthesia, documented treatment preferences are followed postoperatively, and patients are listened to and respected). *[Note: indicators could include those based on direct patient experience]* |
| **11. Multidisciplinary working and care coordination**  Multidisciplinary working, intra and inter-team communication and care co-ordination. | This domain focuses on multidisciplinary working and care-coordination to support the delivery of joined-up care that meets the needs of patients and their caregivers across the whole perioperative care period. It includes a variety of both structural and process indicators.  Structural indicators within this domain could include those relating to:   - **the clarity of overall responsibility for patient care** (such as, whether there is a policy in place that there is a named and documented individual with overall responsibility for patient). - **care pathways, structures and processes in place to support multidisciplinary working and care co-ordination** (such as, whether separate structured pathways are in place for day-case, inpatient and emergency surgeries, and formal defined programmes/pathways or care models for particular patient groups from admission, whether there is timely access available to a range of techniques and services to support care provision – including scanning techniques, reporting by radiologist, biochemistry, haematology, microbiology, and blood bank laboratories, pain management services, inpatient and post-discharge rehabilitation and palliative/pastoral care, whether there is access to a local multidisciplinary and multi-specialty team, with arrangements for the multidisciplinary management of patients with particular needs). - **systems and processes in place to support communication and handovers/transitions of care/escalation** (such as, whether there are systems to support communication within the theatre team about the category of urgency of an emergency, including early warning systems in place with clear escalation processes, and formal handover processes between and within settings). - **protocols and systems in place to support discharge and discharge planning** (such as, whether patients have access to early supported discharge with multidisciplinary team involvement, whether standardised protocols, systems, and criteria are in place governing discharge/ discharge planning)   Process indicators within this domain could include those relating to:   - **adherence to agreed guidelines and pathways for delivery of multidisciplinary perioperative care** (such as, whether plans regarding surgery are communicated appropriately between professionals and teams involved in the patients care, organisation of care involves appropriate multidisciplinary staff, whether agreed handover procedures are followed - using protocols to support communication) - **adherence to agreed discharge protocols and timeliness of discharge** (such as, whether discharge planning processes are initiated and completed in a timely manner, administrative measures such as length of stay in hospital) *[Note: patient reported measures such as functional/mobility status could also potentially be considered here]* |
